# One session of prism adaptation training does not increase immediate engagement in occupational therapy in people with spatial neglect early after stroke: a proof-of-concept study nested within a phase II RCT

**DOI:** 10.1101/2023.07.17.23292600

**Authors:** Matthew Checketts, Ailie Turton, Kate Woodward-Nutt, Verity Longley, Katie Stocking, Andy Vail, Ann Bamford, Audrey Bowen

## Abstract

**Objectives:** Spatial neglect, a debilitating cognitive syndrome and predictor of poor functional outcome, affects attention and awareness after stroke. Early rehabilitation is essential but neglect itself may impede participation in therapy. In a proof-of-concept study nested within an RCT, we investigated whether the oft-reported immediate effects of prism adaptation training (PAT) might enable engagement if introduced at the start of an occupational therapy session.

**Methods:** Early after stroke we video-recorded in-patients carrying out a standardised activity in their first RCT occupational therapy session, before and after PAT (or a control therapy activity). Level of engagement was later scored by a video-rater, experienced in therapy, blind to arm allocation (intervention/control) and whether randomly presented videos were recorded pre-or post-therapy. The rater recorded engagement scores on a 100mm visual analogue scale. Treating therapists also reported, on a 3-point Likert scale, whether or not engagement changed.

**Results:** 49 of the RCT’s 53 patients were recruited (37 PAT, 12 control), 43 of whom consented to be video-recorded. Regression analysis did not suggest improvement in engagement following one session of PAT, using the blinded expert video scoring method: mean difference (95% CI) = −0.5 (−7.4 to 6.4) mm; *p*=0.89). Similarly, post-hoc re-rating of engagement scores (the video-rater viewed paired pre- and post-therapy recordings but remained blind to arm allocation) excluded any material difference in engagement following PAT: mean difference (95% CI) = 1.2 (−2.5 to 4.9) mm; *p*=.52). Impressions of level of engagement provided by the treating occupational therapists also suggested no change: OR (95% CI) = 1.3 (0.13 to 13); *p*=0.81).

**Conclusions:** Despite the need to enable neglect patients to engage in the therapy they are offered, we are confident that a single session of PAT at the start of a therapy session does not enhance immediate engagement in occupational therapy early after stroke. Our study does not address the alternative definition of engagement as a longitudinal, rapport-building process which could meaningfully be explored.

## Introduction and Objectives

Spatial neglect is a common and multifaceted syndrome following stroke that has a profound impact on quality of life. Spatial neglect is sometimes also known as spatial inattention. The manifestation of spatial neglect can vary between people in terms of the area of space and types of stimuli of which the person is unaware. Spatial neglect is most common after right brain damage. Estimates of prevalence vary wildly, however a 2019 study of 88,000 UK stroke survivors estimated incidence among inpatients at around 30% (Hammerbeck et al., 2019), consistent with a systematic review of 41 original research articles (n = 6324) that estimated the prevalence of spatial neglect following stroke at 30% (Esposito et al., 2021).

Spatial neglect is clinically characterised as deficient attention, particularly for contralesional space, and is often accompanied by broader, global attentional dysfunction (Corbetta & Shulman, 2011; Halligan & Robertson, 1999; Rode et al., 2017; Ting et al., 2011). Patients with spatial neglect are likely to experience longer hospital stays and poorer functional outcomes compared to patients without spatial neglect (Chen et al., 2015; Di Monaco et al., 2011; Doron & Rand, 2019; Hammerbeck et al., 2019; Jehkonen et al., 2006; Nijboer et al., 2013). We hypothesised that one factor may be the disruptive effect of the syndrome on full participation in recommended rehabilitation for stroke, regardless of severity or responsiveness to established treatments (Chen et al., 2015; Jehkonen et al., 2006).

Patient engagement in therapy is a key driver of success (Brett et al., 2017). Although therapists routinely appraise engagement informally in clinical practice (Bright et al., 2015; Kortte et al., 2007; Lawton et al., 2016; Lequerica & Kortte, 2010) there is no universally agreed definition. A 2009 survey of occupational therapists in the USA used the following operational definition of engagement: “*a deliberate effort and commitment to working toward the goals of rehabilitation therapy, typically demonstrated through active participation and cooperation*”, p.753 (Lequerica et al., 2009). The concepts of collaboration in goal-setting and therapy work were also highlighted in a review by Bright et al. (2015) who produced a multi-factorial concept of patient engagement, emphasising both the *process* of engagement and *state* of engagement. The latter, engagement state, may vary over time due to internal and external influences and is a potential target for improvement via interventions.

Prism adaptation training (Rossetti et al., 1998) is a straightforward intervention that aims to remediate spatial neglect by way of visuomotor adaptation. PAT does not rely on top-down cognitive processing by its recipients to generate an effect, and so can be carried out with many patients with severe stroke or comorbid anosognosia. PAT has been reported to temporarily improve performance on common cognitive and functional tests for spatial neglect (Lavery & Rowe, 2015; Nys et al., 2008; Priftis et al., 2013; Serino et al., 2009; Shiraishi et al., 2010), however, longer-term effectiveness is yet to be robustly established (Longley et al., 2021). To our knowledge, there are no studies that investigate whether PAT enhances engagement in therapy, but that could be a therapeutic target.

The present study, a proof-of-concept study nested within a randomised controlled feasibility trial of longer term outcome (SPATIAL; Longley et al., 2023), investigated the immediate effect of one session of PAT on patient engagement in occupational therapy. We conceptualised engagement as when a patient is both actively involved in a therapy activity and socially interacting with the therapist.

## Methods

All procedures were approved by a UK NHS research ethics committee (18/YH/0480).

### Recruitment of participants to the proof of concept study

For this proof-of-concept study, we estimated needing 60 patients, allowing 90% power at the 5% significance level to detect a standardised difference of 1.0 standard deviations. Participants for the present study were recruited contemporaneously from stroke service in-patients with spatial neglect who were randomised, with a 3:1 allocation ratio, to the intervention or control arms of SPATIAL; a multicentre, feasibility, randomised controlled trial across the North West of England (Longley et al., 2023). The design, conduct and dissemination of the SPATIAL study was enhanced by active patient involvement from our ‘patient advisory group’ who advised on all aspects of the study.

All participants in SPATIAL received occupational therapy but only the intervention group received PAT, and this was delivered at the start of each therapy session. For the RCT methods and full inclusion and exclusion criteria, see the SPATIAL report (Longley et al., 2023). At the time of consenting to the trial participants were offered the option of also consenting to the present, proof of concept, study which involved agreeing to have the start of their first research therapy session (intervention or control) with their occupational therapist video recorded.

### Prism Adaptation Training in SPATIAL

As described in Longley et al. (2023), the PAT intervention involved up to ninety pointing movements to targets presented in three lateral positions whilst all but the terminal part of the arm movement were concealed by an open-ended box. Pointing movements were performed while 25 prism dioptre (12.5°) prism glasses were worn, with black cardboard blinkers (h=150mm, w=55mm) attached to the sides of the frames to reduce interference from peripheral vision and prevent ambient light from distorting the altered foveal image. Prism direction was determined by neglect side, where patients with neglect of the left side were provided with right-deviating prisms, and patients with neglect of the right side were provided with left-deviating prisms. Full details of PAT in SPATIAL are available in Longley et al. (2023).

### Capturing Engagement: The Visual Scanning Activity

For the purposes of operationalising engagement to investigate instant changes after PAT, we standardised a clinically valid therapy activity for spatial neglect that was a composite of patient behaviours and patient-therapist interactions. This therapy activity and associated engagement scoring sheet was custom-designed for the SPATIAL study and is not intended as a universal research tool to elicit engagement behaviours. These activities were reviewed by our dedicated patient advisory team and local clinicians to ascertain face validity for engagement. Existing measures of engagement are based on self-report and frequent therapeutic interactions between the patient and the same therapist. We were looking for an immediate rather than a longitudinal effect. The issue with self-report with spatial neglect is the common comorbidity of anosognosia, a lack of insight or awareness of difficulties (Grattan et al., 2017). The present study was conducted in a UK National Health Service setting, where different occupational therapy activities are provided by different occupational therapists during the course of the patient’s stay. For this proof of concept study we opted to use a common single activity that was video-recorded before and after an intervention (PAT or control) in the first occupational therapy session post-randomisation. Video recordings were later used for an objective rating of patient engagement.

Our ‘visual scanning activity’ was designed to simulate a practice search task similar to typical process activities used by OTs to help patients to develop strategies to improve their spatial awareness. This activity was selected based on the frequent use of visual scanning in therapy for spatial neglect (Chen et al., 2018). The search activity was not used as an assessment. Instead our intention was that the therapist interacted with the patient, encouraging them, giving suggestions and feedback on performance. Images containing six categories of real-life objects were used, where each category had six exemplars within it. Therapists worked with patients to either search for images of all six of one object category (e.g. “find all the coins”), or a specific exemplar from within a category (e.g. “find the red mug”) on a large, wipe-clean printed poster (see Figure 1) which was secured in the patient’s midline. The poster had a depth of 297mm and a width of 841mm, requiring a wide search involving head and eye movements. Two versions of the visual scanning activity were developed where objects appeared in different locations between versions in order to prevent any learning effects between the two video-recorded time points. The presentation order of the two versions of the visual scanning activities was randomised using a random number generator.

**Figure 1:**
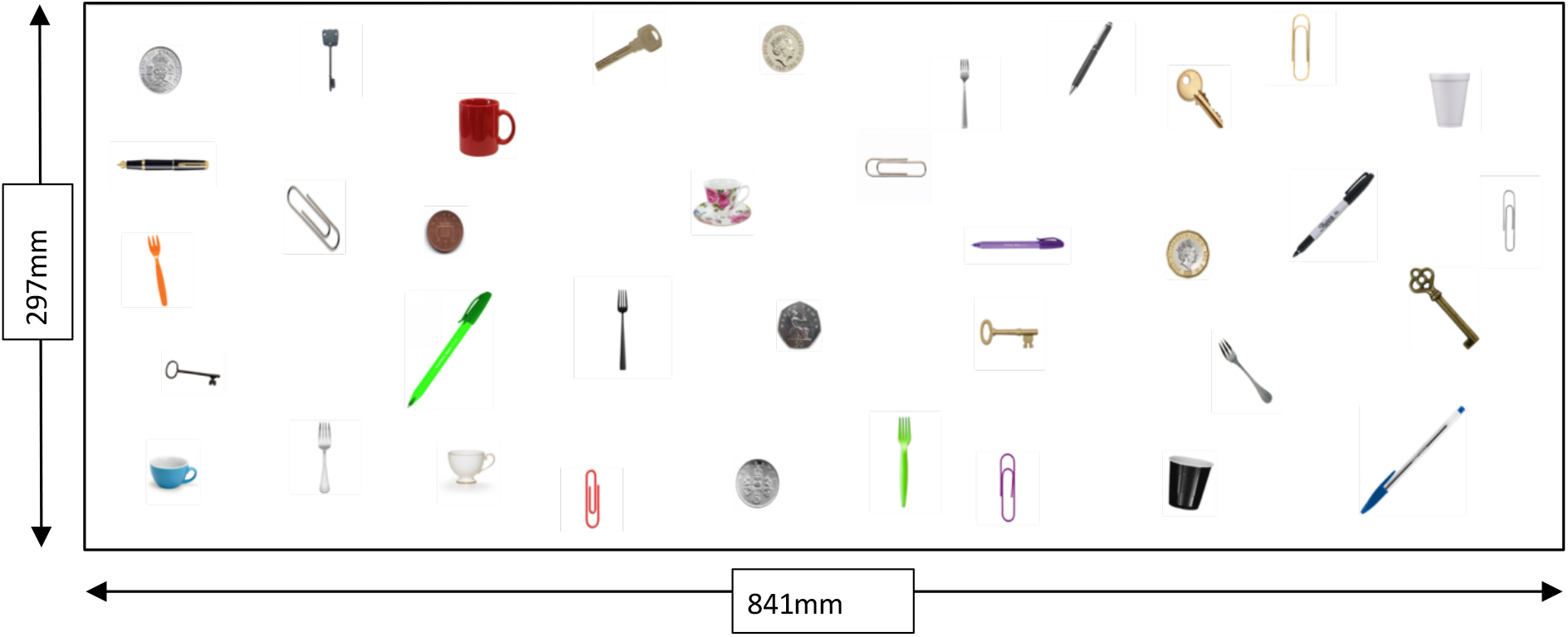
A visual scanning activity developed specifically to simulate an occupational therapy-like interaction, sized to fit on a standard issue adjustable height table. Performance of this activity was video-recorded at two time points per participant.

The visual scanning activity was performed once before and once after 5 minutes of PAT (or a 5 minute standard OT activity in the control arm, e.g. upper limb training) and the entire session (including briefing, activity, and debrief) was video-recorded for each patient. Each patient therefore provided two video clips which were organised into compilations for viewing, after they were individually edited to ensure rater blinding. Videos were recorded on a password-protected and fully encrypted Apple iPad.

The videos were later viewed by a blinded expert rater experienced in OT for stroke, who was guided by a scoring sheet (see Appendix A) containing prompts for observable indicators of engagement. This individual rated engagement on a 100mm visual analogue scale where 0 = no engagement and 100 = best engagement. The scoring sheet, including prompts, was custom-designed for the SPATIAL study and is not intended as a universal research tool for recording patient engagement in therapy. If patients did not give explicit consent to video recording, an unblinded member of the research team observed the therapy session and completed an identical form.

### Video Scoring: Blinded unpaired Viewing

To ensure the video rater was blinded, our *a priori* plan was that videos were randomly organised into compilations of approximately 20 ‘unpaired’ videos e.g. compilations contained either a pre-PAT (or control activity) *or* a post-PAT (or control activity) video from each patient. Videos were separated by a five second interval, allowing the rater to complete the visual analogue scale. The position of the rater’s lines was measured after all the scores within a compilation were collected and recorded in millimetres from 0. This blinded <u>unpaired</u> viewing method was the primary outcome of this study.

### Video Scoring: Blinded paired Viewing

During the study we decided *post-hoc* to compare ratings of engagement, using our purpose-designed scoring method under a second ‘paired’ condition. This was added because the video rater (and the treating OTs) noted difficulty in separating ‘engagement’ from performance (e.g. accuracy, speed, scanning ability) in the visual scanning activity. The ‘paired’ method began only after completion of the unpaired video viewing.

For paired viewing, each compilation of videos contained a pre-PAT (or control activity) *and* a post-PAT (or control activity) video from each patient randomised into the compilation by the trial statistician. The pre- and post-PAT/control video from each patient was presented one after the other (’pre-’ followed by ‘post-’ therapy) to allow within-patient comparison and a patient-specific baseline of therapeutic engagement. Patient videos were separated by a five second interval, allowing the blinded rater to complete the visual analogue scale for each of the pre- and post-therapy videos.

### Treating Occupational Therapists: Unblinded Scoring

At the treatment session, all patients also received a single unblinded rating of engagement from their treating occupational therapist (30 in total across all sites), who received the same prompts as the video rater regarding observable indicators of engagement (see Appendix B). The therapists scored a single tick box from three options: poorer engagement, similar engagement, or improved engagement in the post-PAT (or control) activity compared to the pre-PAT activity.

## Results

### Participant Characteristics

Forty-nine (out of 53) patient participants from the SPATIAL feasibility RCT consented to take part in the proof-of-concept study. Due to the 3:1 randomisation ratio, 37 were from the intervention arm, and 12 from the controls. Of these 49, 43 consented to video recording, and 6 (5 intervention, 1 control) to observation by a researcher who completed the same visual analogue scale used for video rating. However following preliminary data exploration (but before analysis of main effects), we decided to only analyse data from the 43 video-consenting patients rated by a single expert rather than to introduce potential noise by adding in the six ratings from different observing researchers. Characteristics of the 43 patient participants are summarised in table 1.

**Table 1:** Characteristics of SPATIAL patient participants who provided video-recorded proof-of-concept data. NB: The SPATIAL study, where the present proof-of-concept study is embedded, did not routinely collect information on co-morbidities such as cognitive impairment or anosognosia.

|  | **Intervention**<br>**n=32** | **Control**<br>**n=11** | **Whole Cohort**<br>**N=43** |
| --- | --- | --- | --- |
| **Sex** |  |  |  |
| Male n(%) | 19 (59%) | 6 (55%) | 25 (58%) |
| Female n(%) | 13 (41%) | 5 (45%) | 18 (42%) |
| **Age at consent** |  |  |  |
| Mean(SD) | 71 (13.1) | 68 (9.9) | 70 (12.3) |
| Min, Max | 33, 89 | 49, 80 | 33, 89 |
| **Days Post-Stroke on Date of**<br>**Video Recording** |  |  |  |
| Mean (SD) | 20.4 (14.2) | 22.3 (9.6) | 20.9 (13.1) |
| Min, Max | 9, 83 | 8, 42 | 8, 83 |
| **Total NIHSS on Admission** |  |  |  |
| Median | 13 | 11 | 13 |
| IQR | 8.5, 17.0 | 2.0, 16.0 | 7.0, 17.0 |
| Min, Max | 3, 24 | 0, 27 | 0, 27 |
| Missing n(%) | 2 (6%) | 2 (18%) | 4 (9%) |
| **Neglect Side** |  |  |  |
| Left n(%) | 28 (88%) | 9 (82%) | 37 (86%) |
| Right n(%) | 4 (13%) | 2 (18%) | 6 (14%) |

| **Neglect Severity at Baseline -**<br>**OCS Hearts** |  |  |  |
| --- | --- | --- | --- |
| Number Attempted n(%) | 30 (94%) | 6 (55%) | 36 (84%) |
| Median Score | 17 | 9.5 | 16 |
| Min, Max | 3, 47 | 2, 43 | 2, 47 |

Proof of concept participants had a median age of 70, were predominantly male (58%), and of white British ethnicity (95%). They mainly had an ischaemic stroke (79%), on average three weeks previously, affecting their right hemisphere (86%), with a median total stroke severity score (NIHSS) of 13 on admission, resulting in neglect most often of the left side (86%). Several baseline measures of severity of neglect were taken e.g. cancellation tasks (hearts or stars), text reading and a standardised functional assessment of neglect (KF-NAP). Predictably, the proportions rated with mild, moderate and severe neglect varied by test, and were possibly worse on hearts cancellation although only 36/43 (84%) could attempt that test. Of the 36 patients who attempted the hearts cancellation test, 30 exhibited signs of egocentric neglect, four exhibited signs of allocentric neglect, and two presented signs of both subtypes. Overall assessment of neglect severity, by treating occupational therapists based on test scores and clinical observation, was 23% mild, 36% moderate and 40% severe.

### Video Ratings: blinded paired and unpaired

The ladderplots in figure 2 compare the ‘pre’ and ‘post’ scores from the blinded video rater per participant according to study arm and mode of video viewing. These suggest there is less variability in scoring for the paired viewing, compared to the unpaired and that the video rater’s change scores were very small, particularly for paired viewing.

**Figure 2:**
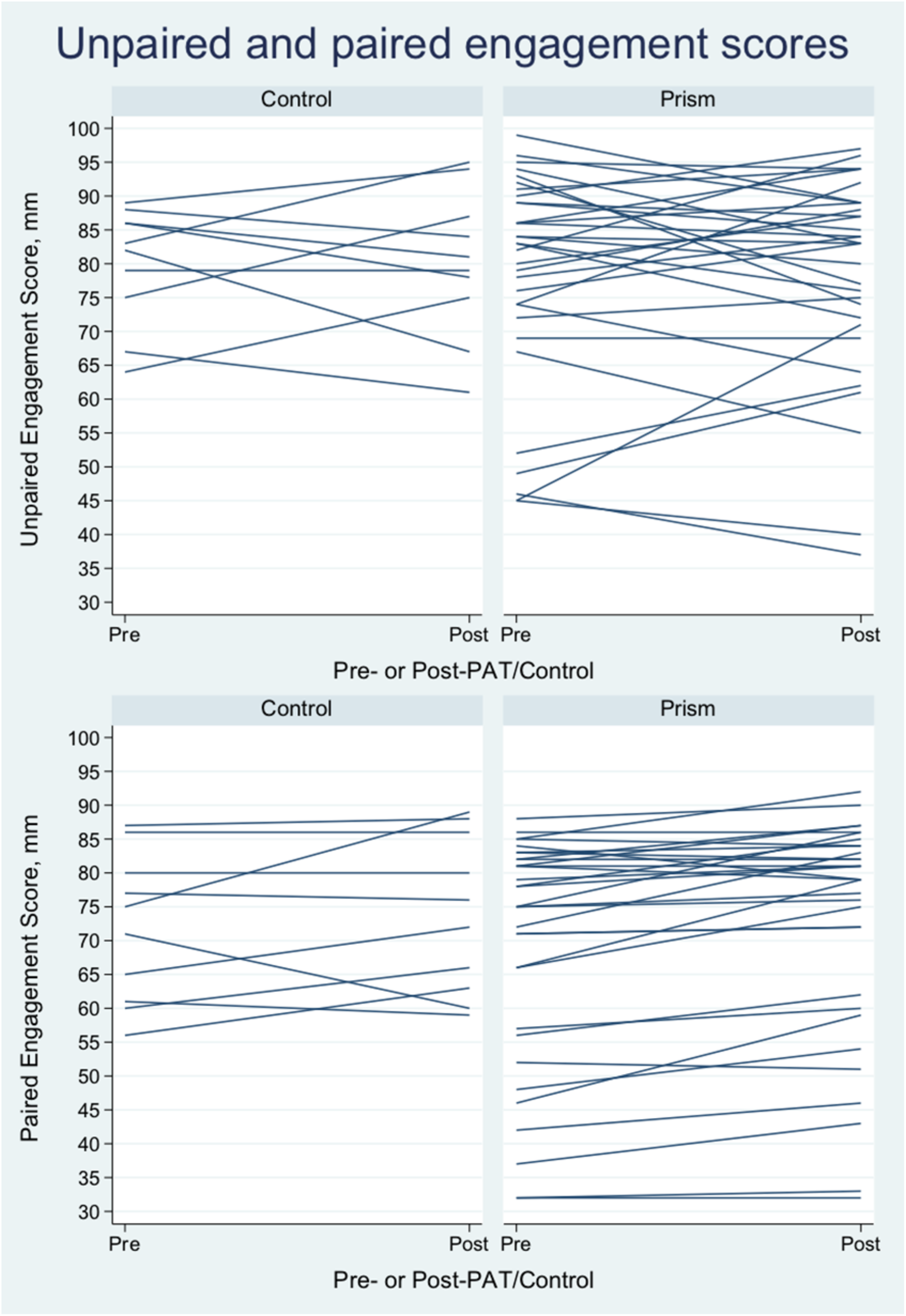
Ladderplots displaying the pre- and post-therapy engagement scores from the blinded video rater, by arm, for the unpaired viewing (top) and paired viewing (bottom).

Mean blinded unpaired engagement scores on a 100mm visual analogue scale are summarised in table 2. Participants in the prism arm were rated as having a slightly smaller engagement change scores on average than control participants, although the variability is large within each group. Linear regression analysis of the unpaired ‘pre’ and ‘post’ engagement scores found no evidence of improvement in engagement following prism adaptation training, recording a mean difference (95% CI) of −0.5 (−7.4 to 6.4) mm; p=0.89 with quite a wide confidence interval.

**Table 2:** Blinded unpaired video engagement scores in millimetres (100mm visual analogue scale). *Due to corrupted video data, one control patient did not receive a ‘post’ therapy engagement score and was thus removed.

| **Arm** | **Unpaired 'Pre'**<br>**(mean, SD)** | **Unpaired 'Post'**<br>**(mean, SD)** | **Unpaired Change**<br>**(mean, SD)** |
| --- | --- | --- | --- |
| PRISM (n=32) | 78.4 (15.7) | 78.5 (15.1) | 0.1 (10.2) |
| CONTROL (n=10)* | 79.9 (8.7) | 80.1 (10.8) | 0.2 (9.4) |

Mean blinded paired engagement scores on a 100mm visual analogue scale are summarised in table 3. As anticipated, paired video viewing reduced the variation (SD) of the change scores. Participants in the prism arm on average were rated to have larger change scores than those in the control arm, but these differences were small, i.e. a few millimetres. Linear regression of the paired ‘pre’ and ‘post’ engagement scores found no evidence of improvement in engagement after prism adaptation training, recording a mean difference (95%) of 1.2 (−2.5 to 4.9) mm; *p*=0.52. The reduced variation led to a tighter confidence interval, ruling out a true difference (determined *a priori*) of 10mm or more between arms.

**Table 3:** Blinded paired video engagement scores in millimetres (100mm visual analogue scale). *Due to spoiled video data, one control patient did not receive a ‘post’ therapy engagement score and was thus removed.

| **Arm** | **Paired 'Pre'**<br>**(mean, SD)** | **Paired 'Post'**<br>**(mean, SD)** | **Paired Change**<br>**(mean, SD)** |
| --- | --- | --- | --- |
| PRISM (n=32) | 69.0 (17.1) | 72.5 (16.7) | 3.4 (4.4) |
| CONTROL (n=10)* | 71.8 (11.0) | 73.9 (11.6) | 2.1 (6.8) |

### Engagement Ratings – Unblinded Treating Therapist Ratings

Figure 3 displays the unblinded therapist ratings by arm. Three participants were rated as having poorer engagement after therapy compared to before (two in the prism arm, one in the control arm). Forty-one participants were rated as exhibiting no change in engagement (31 in the prism arm, 10 in the control arm) and 5 participants were rated as exhibiting improved engagement (4 in the prism arm, 1 in the control arm). Binary logistic regression analysis revealed no evidence of improvement in engagement following PAT, recording an odds ratio (95% CI) of 1.3 (0.1 to 13.2); *p*=0.81. These findings are consistent with those generated by the blinded video viewing procedures, above.

**Figure 3:**
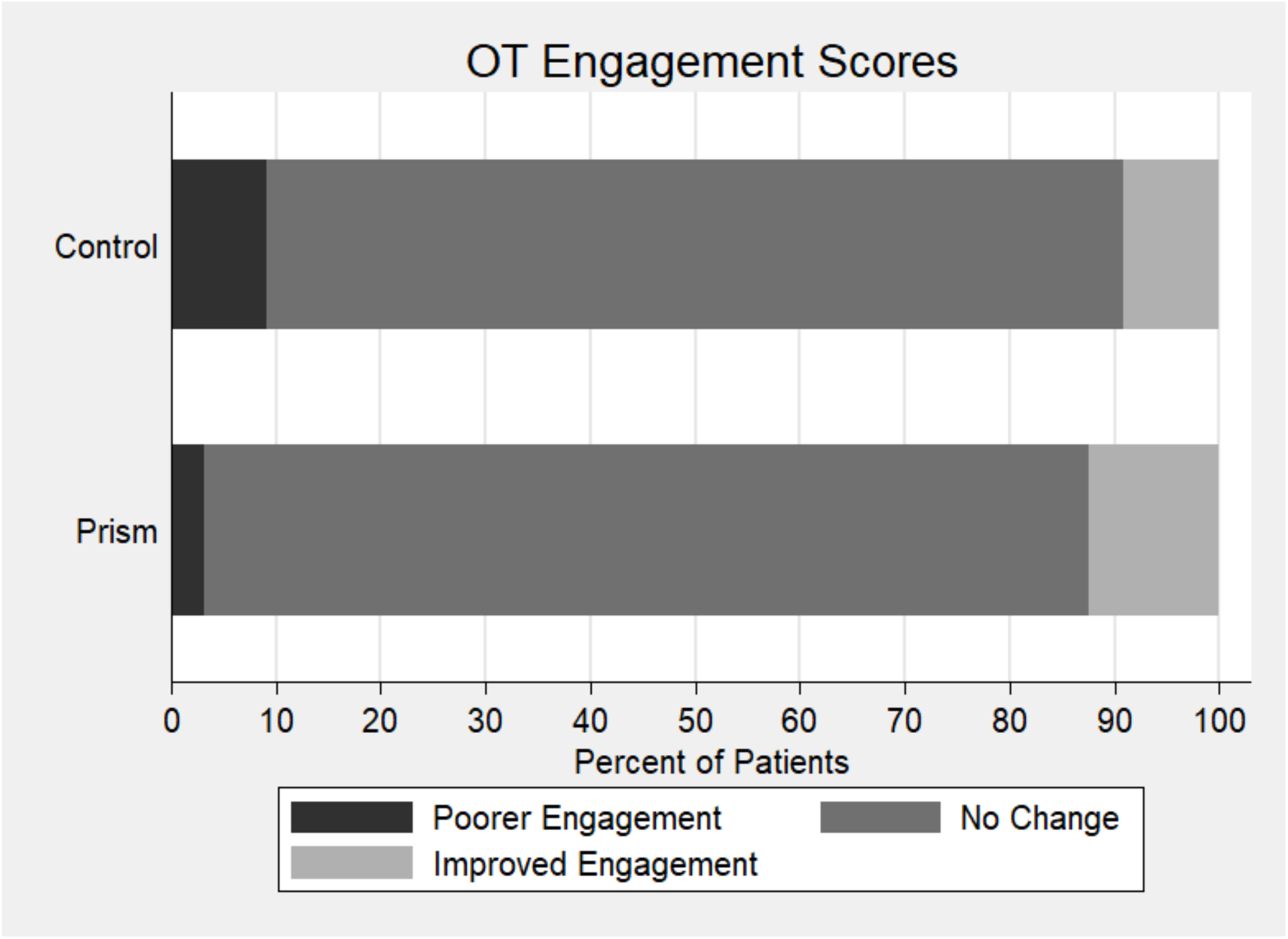
Unblinded treating therapist engagement ratings by arm.

## Discussion

### Summary of results

By nesting a proof-of-concept study within a feasibility, randomised controlled trial (Longley et al., 2023) we explored whether a single dose of PAT for spatial neglect immediately improves patient engagement within that session of occupational therapy. Our novel method to objectively measure immediate engagement, created specifically for this study rather than as a standalone measure of the broader concept of therapeutic engagement, found that immediate engagement was not improved within one occupational therapy session incorporating PAT. Despite not reaching our target sample size of 60, the study was sufficiently powered to rule out what may be considered a minimally important difference of 5mm on the VAS. This finding is consistent across both unpaired and paired video viewing methods (from a blinded single rater, thereby removing concerns about inter-rater reliability) and concurs with ecologically valid ratings of no change in engagement from unblinded treating occupational therapists. However, before concluding there is no effect of PAT on engagement, it is important to consider alternative explanations for this consistent finding and the strengths and limitations of our study.

### Measuring immediate engagement: how good was our novel method?

The key strengths of the study were the novelty of our methods, enhanced validity through patient involvement as research partners and input from occupational therapists, and our efforts to minimise bias. Furthermore, all of our results (blinded paired, unpaired, and unblinded therapists’ scores) simultaneously suggest that one session of PAT does not enhance engagement immediately. The initial unpaired video scoring method (a standardised visual scanning activity) was designed to ensure full blinding of the video rater with respect to allocation (intervention/control) and time point (pre-or post-therapy). According to the rater, the concealment was maintained throughout unpaired viewing. An unforeseen consequence was that this made it difficult to separate ‘engagement’ from pure ‘performance’ on the visual search task (e.g., accuracy, speed, scanning ability). Returning to conceptualisations of engagement, Bright et al. (2015) highlight the ‘state’ of engagement, which we attempted to capture in video recordings, but also the longitudinal ‘process’ of engagement. It could be argued that the process of establishing engagement could occur in parts of the task not always captured on video, e.g. during task set-up and instructions, which occasionally contained dialogue that would unblind the video rater and was thus eliminated on a case-by-case basis from the processed videos. Whilst no patient was rated at 100% (’full engagement’) on the visual analogue scales, some of the pre-therapy scores are above the 50% and even the 75% level particularly in the blinded unpaired condition. Whilst these levels of engagement do indeed leave room for improvement, it is possible that this patient group with spatial neglect does not have the severest impairments of engagement that we were anticipating.

The paired video scoring method was added to the study following scoring of the unpaired videos and feedback from the rater (and treating therapists) but before analyses were conducted. In paired viewing, the pre- and post-therapy videos of each patient were presented together allowing for a within-patient comparison of engagement change (although importantly the rater was still blinded to allocation to PAT/control). This method, according to the video rater, made it easier to rate the state of engagement independently, without any performance confound. This is reflected in our results, where the variability for paired scoring is markedly less than in unpaired viewing. As with the unpaired viewing the rater did not use the maximum value of 100% engagement on the visual analogue scale, although any change in engagement score is minute when the millimetre scale of the visual analogue scale is considered. Compared to the unpaired pre-therapy scores, the paired scores recorded for the pre-therapy videos tended to be lower. One potential explanation for this is that the rater had an expectation of poor engagement pre-therapy, and of response to therapy, which is exactly why we initially designed the study to examine unpaired blinded data and why these remain our primary analysis. Nonetheless the paired data provide a useful sensitivity analysis and show consistency in our finding that one session of PAT does not immediately enhance engagement in OT.

The videos were all recorded in the first OT session within the SPATIAL feasibility RCT. One might question whether patients who were on average only three weeks post stroke would exhibit changes in engagement observable on video by a rater not involved in their care. For example, there may be an optimal later time point post-stroke where patients possess sufficient cognitive resources to engage in therapy or for this to be observable based on a short viewing. It could also be the case that patients who met the inclusion criteria for SPATIAL were those with enough cognitive capacity to participate in occupational therapy (and a feasibility trial), and those with the most impaired engagement may have been excluded. With the support of our patient advisory group we encouraged the therapists to be as inclusive as possible and people with severe stroke and severe neglect were included, including those requiring tilt-in-space chairs. However, we cannot rule out the possibility of exclusions based on perceived suitability for participation by recruiting staff.

### How useful was the inclusion of engagement ratings from the treating occupational therapists?

Another strength of the study was the triangulation of data. In addition to the objective blinded (unpaired and paired) video ratings the NHS occupational therapists delivering the interventions were also asked about their impressions of their patients’ engagement immediately after the first OT session. The treating therapists were given three choices: poorer engagement, similar engagement, or better engagement - in the post-therapy visual scanning activity session compared with the pre-therapy visual scanning activity session. The majority of scores obtained in this way indicated no change in engagement which concurs with the findings from the blinded video scoring methods. Of course, one could argue against our conceptualisation of engagement, which we freely accept took a snapshot approach, and suggest that the ‘process’ of engagement development between patient and therapist and the resulting ‘state’ of engagement should be considered over a period of time and with more naturalistic observation or qualitative interview. The downside of these alternative approaches would be that factors such as variability in the interpersonal skills of different therapists would make it difficult to measure the effects of PAT. Notwithstanding possible limitations of our methods, they had many strengths and, in our experience, we would have expected to see some indication of an increase in engagement had it been present. In fact, the limitation is that we succeeded in focusing objectively on immediate engagement in a standardised task and it remains to be established whether the much broader concept of therapeutic engagement is affected by PAT, if indeed that broader concept could be reliably captured.

### Is one session of PAT an insufficient dose?

Several studies have suggested PAT rapidly ameliorates spatial neglect in stroke patients. A recent case study conducted by Abdou et al. (2020) claimed that a single PAT session significantly improved sitting balance as measured by electromyography of the patient’s trunk muscles and a pressure mat. In terms of neural mechanisms of PAT effects, Crottaz-Herbette et al. (2017) reported that a single exposure to prisms in patients with spatial neglect significantly enhanced compensatory mechanisms situated in the intact left hemisphere and rapidly improved neglect behaviours. These findings require testing in adequately powered randomised controlled trials but justify our decision to look for an immediate effect on engagement following the first dose of PAT.

There is some evidence to suggest that PAT effects observed in patient participants extend beyond trained visuomotor and spatial attention tasks to wheelchair driving, navigation, visuo-verbal tasks, and even beyond the visual domain in tactile or haptic tasks, and in auditory extinction (Jacquin-Courtois et al., 2013). Whilst this is encouraging in terms of potential expansion to engagement in therapy, there is no strong evidence that these effects are elicited after a single exposure to PAT. Indeed, the findings of the present study suggest that PAT did not increase engagement. We cannot say whether a series of PAT sessions would have had a greater effect. However, in our separate feasibility trial paper we report an absence of any benefits on a range of outcome measures after three weeks of PAT, consistent with the findings in the current paper.

Moreover, some evidence suggests that PAT is more specific in terms of neural adaptation than previously claimed, and thus may not be a useful intervention for the broader cognitive and emotional processes likely to be involved in therapeutic engagement. A 2021 review of functional magnetic resonance imaging studies found that, in healthy participants, PAT is associated with activity in the posterior parietal lobe and the cerebellum, whereas in participants with neglect following right brain damage, there is a lack of a consistent pattern of neural activity, possibly as a result of reduced connectivity and reliance on alternative circuitry to perform spatial tasks (Boukrina & Chen, 2021). Milner and Goodale (Goodale & Milner, 1992; Milner & Goodale, 2012) proposed that visuospatial processing is composed of two routes: the allocentric ventral stream, coding for the recognition and identification of objects in space and located in the ventral temporal cortex, and the egocentric dorsal stream, coding for object location and visually guided behaviours that help to physically locate these objects and located in the superior parietal lobule and inferior parietal sulcus. A case study by Mancuso et al. (2018) reported that PAT reduced symptoms of egocentric neglect (i.e. neglect relative to bodily midline), but not allocentric neglect (i.e. neglect relative to the midline of individual objects).

Furthermore, a review by Striemer & Danckert (2010) concluded that the beneficial effects of PAT are a result of direct modulation of the dorsal stream (and thereby egocentric processing). These beneficial effects of PAT may also rely on successful neural communication between the dorsal and ventral streams, which is precluded by lesions of the inferior parietal lobe and/or the temporal gyrus - areas which are often damaged in patients with spatial neglect (Corbetta & Shulman, 2011; Mort et al., 2003). It is noteworthy here that 30 of the patients in the present study exhibited signs of egocentric neglect on the hearts cancellation test and still did not appear to exhibit enhanced engagement following PAT.

### Does our conceptualisation of engagement affect how we study it?

Engagement in therapy, for example in occupational therapy, is a factor considered likely to enhance or impede recovery from stroke. Engagement has a broad conceptualisation that presents challenges for quantitative investigations (Brett et al., 2017; Kennedy & Davis, 2017). For example, the definition of engagement from Bright et al. (2015) encompassed the quality of the relationship between the patient and therapist/service, perceived usefulness of treatment, co-construction of rapport, as well as some ‘observable’ constructs such as willingness to participate, persistence and determination, and assertion of personal identity. Lawton et al.’s concept of therapeutic alliance extends some of these factors to factors outside of the therapy setting, including ‘optimal’ involvement of friends and family in recovery, and more tangible system drivers that include organisational and financial constraints (Lawton et al., 2016). The approach taken by Lequerica et al. (2009) further expands the focus of ‘engagement in therapy’. The authors identified several barriers (e.g. depressed mood, working memory impairment, aversion to correction, confusion, decreased alertness) and facilitators (e.g. support for meaningful activities, rapport, patient control, flexibility) related to patient engagement. Whilst these findings may have validity in the population they recruited from, they don’t fit our conceptualisation or inform our measurement of patient engagement as an immediate outcome of PAT. We acknowledge that we focused on a robust investigation of just one aspect of engagement, although we suggest our findings provide novel evidence that warrants caution about PAT for neglect early after stroke. These findings, and the absence of benefit from the outcomes measured in our feasibility trial elsewhere, have been sufficient to halt our plans to proceed to a definitive trial of early PAT and have sent us in search of alternative interventions for this patient population.

We consider the probable range of cognitive impairments present in our sample of stroke survivors to be a particular strength of the larger SPATIAL study, given that these patients are often overlooked in clinical research. However, it has been consistently reported that stroke survivors who have spatial neglect often present with a mixed neurocognitive profile (Ting et al., 2011), possibly resulting from large lesions that disrupt several cognitive processes (Li & Malhotra, 2015; Verdon et al., 2010) which may hinder engagement in therapy. These different profiles may result in different requirements for measuring and improving engagement. As mentioned previously, some measures of therapeutic engagement exist, although we consider these to be inappropriate for the present study because they rely on self-report (Kortte et al., 2007; Lenze et al., 2004), or because they have been developed for use in different clinical populations (e.g. dementia (Cohen-Mansfield et al., 2011), mental health (Hall et al., 2001)), or in different therapies (e.g. Speech and Language Therapy (Simmons-Mackie & Damico, 2009) or group therapy (Roy et al., 2012)).

Finally, previous studies that sought to define the concept of engagement tended to utilise qualitative rather than quantitative methodology. This has ramifications for clinical research and ultimately practice, since interventions cannot be developed or tested without a set of definitions and outcomes to guide researchers. Efforts should be made to synthesise the qualitative data available with a view to transferring this rich knowledge to quantitative investigation to further investigate the effect of interventions on engagement and ultimately on outcomes.

### Strengths and limitations

In addition to its strengths this study has limitations. First, we took care in our attempt to measure the immediate effect of PAT on engagement in occupational therapy, a process aided by a dedicated SPATIAL advisory group of stroke survivors. However, we did not measure the immediate effect of PAT on established neuropsychological and functional measures of spatial neglect, nor did we measure whether individual patients adapted to the prisms post-treatment. However, we did stipulate a minimum amount of PAT that patients should receive, set at either 5 minutes or 90 pointing movements with prisms on (whichever is earliest). Our feasibility trial outcomes suggested good adherence to the treatment protocol. This intensity of PAT is surmised to be a broad enough criterion to allow all patients to adapt properly and adaptation was demonstrated in our previous work with a similar population (Turton et al., 2010).

Secondly, this proof-of-concept study was nested within a randomised controlled trial study and benefited from the reduction of bias however there was a difference in inter-video activities between the two study arms. The participants in the prism arm received five minutes of PAT, whereas the control group received five minutes of a “control intervention”. We deliberately did not stipulate what the control activities could be as we wanted it to be usual occupational therapy e.g., some patients received upper limb training, additional visual scanning tasks, and formal neuropsychological testing. It could be the case that patients in the control arm received stronger social primers for ‘engagement’, particularly given that PAT is generally a quiet activity, and OT interventions tend to involve more conversation.

Thirdly, the activity we designed to simulate OT for the purposes of video recording may not have elicited engagement behaviours as we intended. The visual scanning activity was designed to be a collaborative and active therapy task, whereas in reality some patients and therapists may have treated it more like an assessment of cognition and function. Indeed, the video rater noted that some therapists were silent during the visual scanning activity and may not have provided patients with the opportunity to demonstrate their engagement.

Fourthly, we did not systematically capture which co-interventions individual patients might have been receiving. In the UK, stroke patients are usually offered physiotherapy, nursing and medical care and to a lesser extent speech and language therapy and psychology. Differential combinations of input from these services may have influenced the state of engagement. However, the randomisation procedure stratified by site should have minimised this. Additionally, whilst a strength of the SPATIAL study was that we recruited from a large range of sites increasing generalisability of our findings, the between site differences in service provision might be viewed as a limitation.

Finally, individual differences may be a confound in this study such as different stroke aetiologies (lesion location and extent, single or multiple strokes, left or right neglect) or co-morbidities (e.g. visual field defects) influencing engagement behaviours and the effects of PAT. However, the randomisation should have balanced these between the prism and control arms. Likewise, individual patient characteristics may make it difficult to objectively rate engagement. For example, some patients may use humour at different rates or for different reasons or may have physical impairments such as postural instability that make them appear less engaged.

## Conclusions

There is no evidence that a single session of PAT enhanced immediate engagement in occupational therapy. In fact, our proof-of-concept study ruled out any meaningful effect on engagement (as defined by us). This conclusion is strengthened by triangulating different measurements: blinded (unpaired) ratings of videos of a study-specific standardised therapy activity, ratings of paired videos, and the perceptions of the therapists providing the interventions However, a broader conceptualisation of engagement, as a longitudinal, rapport-building process involving interaction with both the therapist and the therapy activity, might not be captured using our snapshot method, and may not be a straightforward target for the PAT intervention. Questions remain regarding whether a larger dose of PAT could increase engagement, or whether PAT needs to be delivered at a later time post stroke or in a more specific subset of patients with neglect, and whether the broadest conceptualisation of engagement in therapy can be quantified and manipulated for improving outcomes in rehabilitation research.

## Data Availability

All data produced in the present study are available upon reasonable request to the authors.

## Acknowledgements

This independent research was funded by the National Institute for Health Research (NIHR) under its Research for Patient Benefit (RfPB) Programme (Grant Reference Number PB-PG-0816-20016). The views expressed are those of the author(s) and not necessarily those of the NHS, the NIHR or the Department of Health and Social Care. Authors also acknowledge funding from The University of Manchester Research Impact Scholarship (MC) and Stroke Association, UK (AB; grant number TSA LECT 2015/01 – SCOPE: Strategies to COPE with cognitive difficulties after stroke). Funders had no role in study design, execution, analysis or results interpretation.

The authors extend thanks to patient participants, therapy and research staff, our dedicated patient advisory group, members of the trial management group, members of the independent trial steering committee, and to Jessica Haigh for support in video data processing.

The authors declare no conflicts of interest.

---

**Appendix A:**
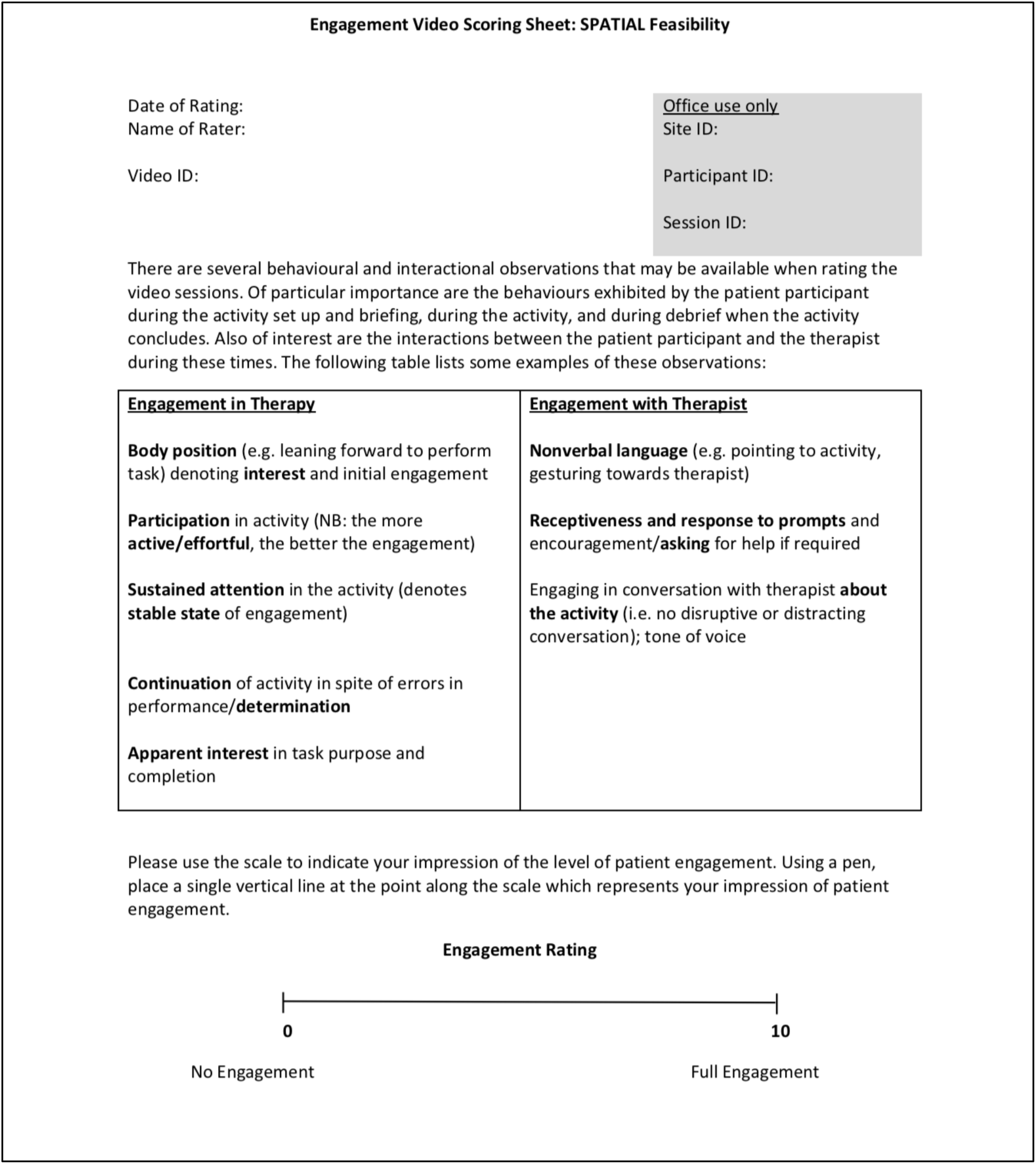
Video rater engagement scoring sheet

**Appendix B:**
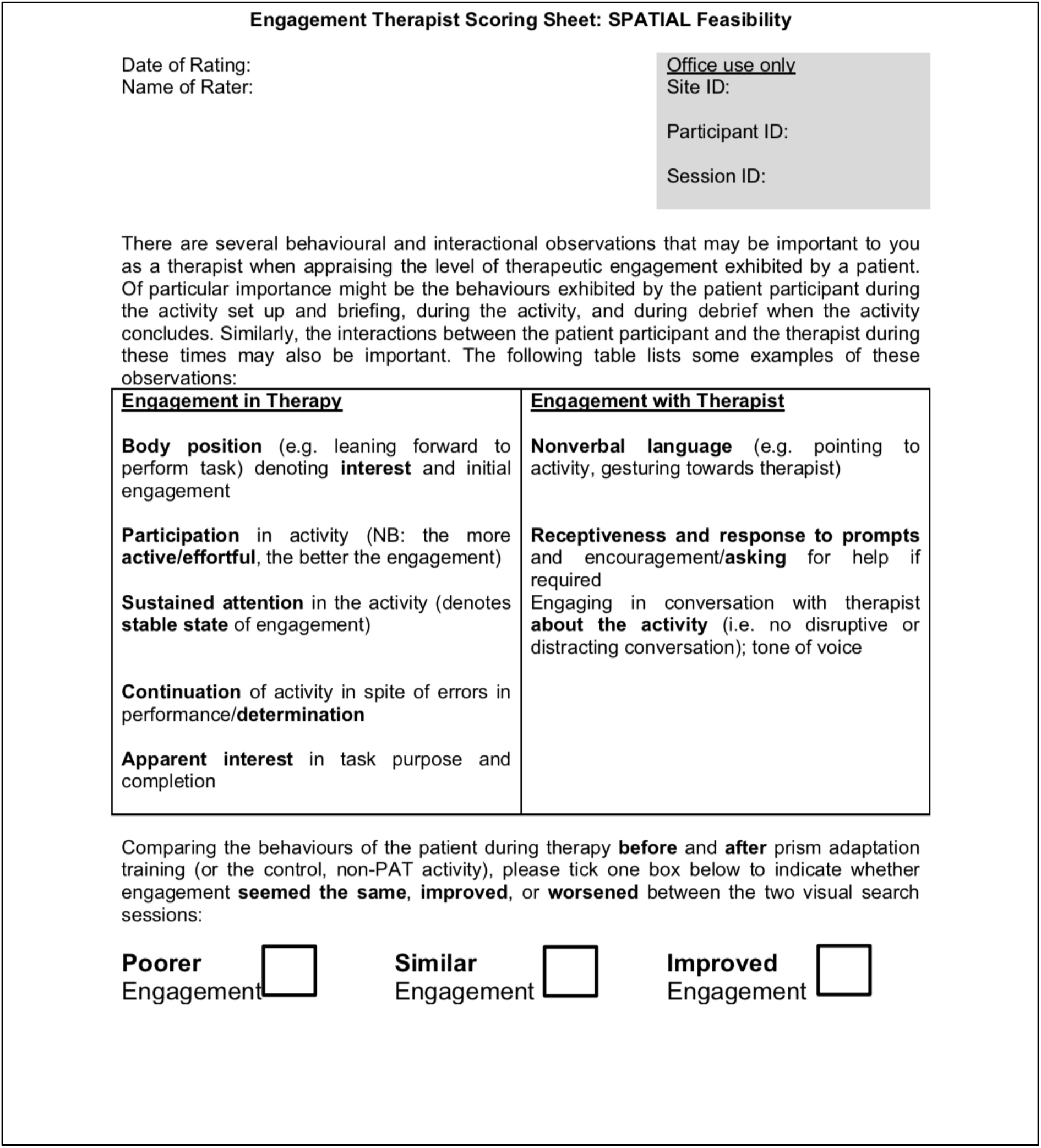
Treating OT engagement scoring sheet

